# The Effect of chlorhexidine mouthwash on the color stability of three types of dental resin composites

**DOI:** 10.1101/2025.05.23.25326136

**Authors:** Elnaz Shafigh, farnam esfahani, siavash abdollahpoor, navid shafigh

## Abstract

**Objective:** The successful application of composite resin restoration largely depends on their color stability, and discoloration is still a problem in composite resin restorations due to the desired instability. Chlorhexidine is one of the most widely used mouthwashes. The most important side effect of chlorhexidine is tooth discoloration. Since there has been no study on the effect of chlorhexidine application on microhybrid, nan and packable composites, the present study will be performed by spectrophotometric laboratory method to measure the staining of these composites due to contact with 0.2% chlorhexidine.

**Materials and Methods:** Nano filler, Packable and Micro hybrid composites are choose for our study for testing 35 samples of each composite were prepared in 2 mm in 1 cm disks. and then all three groups (n=105 samples) are exposed to 0.2% chlorhexidine for one minute every day for 2 weeks. Finally, discoloration will be investigated in each group.

**Results:** CHX provided the significant color change in all of the 3 type of composite groups (P<0.05). Data analyses are seen in the table one two and three. Between 3 type of composite z350 micro hybrid has a best color stability and nanofiller z350 is more prone to discoloration in comparison of Packable P60.

**Conclusion:** According to the results of the present study and related limitations, it can be concluded that micro hybrid composite is the best type of composite in tooth restoration of people who have to use chlorhexidine for a long time.

## Introduction

Discoloration of composite restorations due to contact with various intraoral materials has been an important issue in recent years. Numerous materials have been analyzed as staining agents, typically beverages and mouthwashes. Discoloration of composite restorations is one of the common reasons for replacing these restorations, which can occur due to internal and external factors. ^12^

Mouthwashes with antibacterial effects inhibit the bacterial growth in the mouth and teeth. Chlorhexidine is one of the most widely used mouthwashes, which has many applications considering its ability to inhibit dental plaque and prevent decay of smooth tooth surfaces. The anti-plaque effect of chlorhexidine is due to the special nature of its chlorhexidine molecule, which guarantees the stable antimicrobial effect of this substance on the tooth surface through bactericidal and bacteriostatic effects. ^3^

The most important side effect of chlorhexidine is tooth discoloration. This discoloration occurs due to the cationic nature of the chlorhexidine molecule and local deposits due to the reaction between chlorhexidine adhering to the tooth surface and chromogens in foods and beverages .^4^

Discoloration of dental restorations can be detected both visually and using laboratory equipment. Instrumental methods are more reliable than ocular methods .^5^ Among these methods, spectrophotometry is the most accurate method to detect discoloration of dental restorations.^6^ Although there are many similarities between various staining measurement methods and spectrophotometry, there are still many differences that can be used in choosing the best staining measurement method in dental restorations.^7^ Spectrophotometry is an extremely powerful method that can show more accurate and in-depth measurements than other colorimetric methods such as spectral data. Therefore, this method is used for accurate measurements in research and development or laboratory comparisons. In contrast, other colorimetric methods are simpler and are mostly used for production and quality control purposes in factories. ^8^

Farhan and Gregorius, W.C., et al show that all of acrylic dentures have color change in discoloring materials. ^910^

Since there has been no study on the effect of chlorhexidine application on micro hybrid, Nano filled and packable composites, the present study will be performed by spectrophotometric laboratory method to measure the staining of these composites due to contact with 0.2% chlorhexidine.

## Materials and Methods

Nano filler resin composite (z350) (3M, USA), P60 (3M, USA) (Packable) and Micro hybrid resin composite (Z250) (3M, USA) were treated with chlorhexidine mouthwashes(0/2 % CHX, irsha,iran).

For testing 35 samples of each composite were prepared in10 mm in 2 mm in 1 cm disks. 105 cylindrical disc specimens (35 specimens for each material) were prepared sing Teflon molds (diameter 10 mm and thickness 2 mm). A transparent polyester strip band (Mylar strip; SS White Co. Philadelphia, PA, USA) and glass plate were used in the application of light pressure to remove excess material and obtain a smooth surface. The specimens were polymerized using an LED device (woodpecker china) with a light intensity of 600 mW/cm^2^. The distance between the light source and the sample was standardized using a 1 mm transparent polyester strip band. After polishing with a Super-Snap Rainbow Technique Kit (Shofu, Inc., Kyoto, Japan) and One Gloss Polishing Kit (Shofu, Inc., Kyoto, Japan .^11^

To perform spectrophotometry, the samples were measured using a Xrite I1-Pro portable spectrophotometer with radiation / measurement geometry of 45^°^ / 0^°^. The colorimetry test was performed in a light and color laboratory under the supervision of a specialist. for the easier method to analyze each 5 pieces of disks were analyzed together and mean of them are shown in the table.

The initial color values (T□) of the samples were measured using an digital spectrophotometer operated in “shade of restoration” mode. The device was first calibrated using a white calibration plate in accordance with the manufacturer’s recommendations, and then measurements were performed. The device was recalibrated after every 15samples measured. The measurements were repeated three times for each sample, and mean values were subjected to analysis. The samples were randomly divided into five groups before immersion in mouthwash solutions. The number of samples to be studied was estimated with a power of 90% and an assumed significance level of .05 implemented in the estimation. A sample size of n=35 was determined.

Group 1: 35 samples nanohybrid composite

Group 2 : 35 samples microhybrid composite

Group 3 : 35 samples pachable composite

Group 4 : five disk-shaped samples from all three groups (n=15 samples) are exposed to normal saline as a positive control

Group 6 : 35 ceramic samples as a negative control

all three groups (n=105 samples) are exposed to 0.2% chlorhexidine for one minute every day for 2 month.

Following, the samples washed with distilled water, and dried with a sponge, after which the color measurement procedures were repeated (T□). These procedures are summarized in <u>Figure 1</u>. The changes in color values (ΔE□ □) between T□ and T□ were calculated using the CIEDE2000 color system and the following formula ΔE□ □:

((ΔL/kLSL)^2^ +ΔC/kCsC)^2^+(ΔH/kHsH)^2^ +Rt(ΔC/kCsC) (ΔH/kHsH)) Where ΔL, ΔC, and ΔH represent the differences in lightness, chroma, and hue between the initial and subsequent color measurements, respectively. SL, SC, and SH are the functions of weight, incorporated into the formula to eliminate the irregularities observed in the CIE L*a*b system; they refer to brightness, color density, and hue, respectively (weighting functions0^12^ These values reflect the total color difference among L*, a*, and b* values in two different coordinates. After examining the samples of similar experiments, the average of 35 samples for each of the studied materials (three groups: a total of 105 disk-shaped samples) and 5 positive control samples for each of the studied materials (A total of 3 composite groups: 15 disk samples) and 15 negative control ceramic samples were selected. A total of 130 laboratory samples will be tested on the tested disks.

### Statistical analysis method

All statistical analyses were performed using the Statistical Package for the Social Sciences (SPSS Inc., Chicago, IL, USA). three-way ANOVA in SPSS ver. 25. In the event of a significant difference between the materials, Duncan’s post hoc multiple comparison test was performed. The level of statistical significance was set at p=0.05.

**Table 1.** Compositions of the composite resin materials.

| Restorative materials | Type | Filler Composition | (%) Weight, Volume | Particle size | Manufacturer |
| --- | --- | --- | --- | --- | --- |
| Z350 | Nano filler resin composite | The fillers are a combination of non-agglomerated/non-aggregated 20nm silica filler, non-agglomerated/non-aggregated 4 to 11nm zirconia filler, and aggregated zirconia/silica cluster filler | The inorganic filler loading is about 72.5% by weight (55.6% by volume) for the Translucent shades and 78.5% by weight (63.3% by volume) for all other shades. | The fillers are a combination of non-agglomerated/non-aggregated 20nm silica filler, non-agglomerated/non-aggregated 4 to 11nm zirconia filler, and aggregated zirconia/silica cluster filler (comprised of 20nm silica and 4 to 11nm zirconia particles). | 3M (USA) |
| P60 | Packable resin composite | The filler in Filtek P60 restorative is zirconia/silica | The inorganic filler loading is 61% by volume (without silane treatment) | particle size range of 0.01 to 3.5 μm. Filtek P60 restorative contains BIS-GMA, UDMA, and BIS-EMA resins. | 3M (USA) |
| Z250 | Micro hybrid resin composite | The filler in Filtek Z250 restorative is zirconia/silica | The inorganic filler loading is 60% by volume (without silane treatment) | article size range of 0.01 to 3.5 μm. Filtek Z250 restorative contains BIS-GMA, UDMA, and BIS-EMA resins. | 3M (USA) |

**Table 2.**
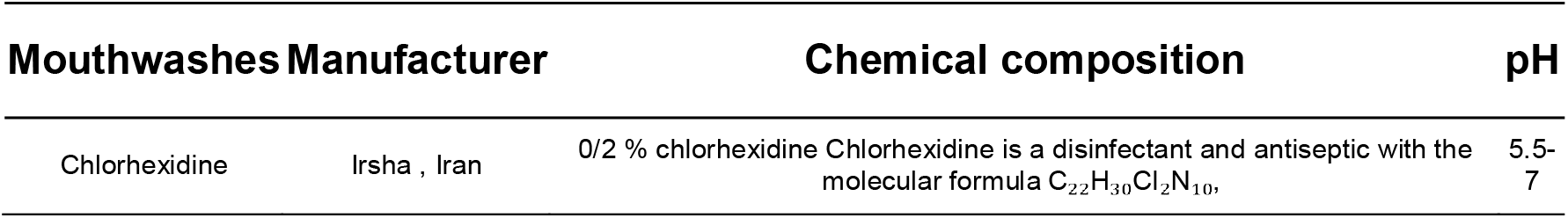
Chemical compositions of the mouthwashes.

## Result

CHX provided the significant color change in all of the 3 type of composite groups (P<0.05) Data analyses are seen in the diagram one –six. Between 3 type of composite z350 micro hybrid has a best color stability and nanofiller z350 is more prone to discoloration in comparison of Packable P60.

Between 3 types of composite the color change isn’t significant and they don’t have a difference with each other. The diagrams 1 to 6 show the color CIELAB of each composite before and after CHX exposure.

**Diagram 1.**
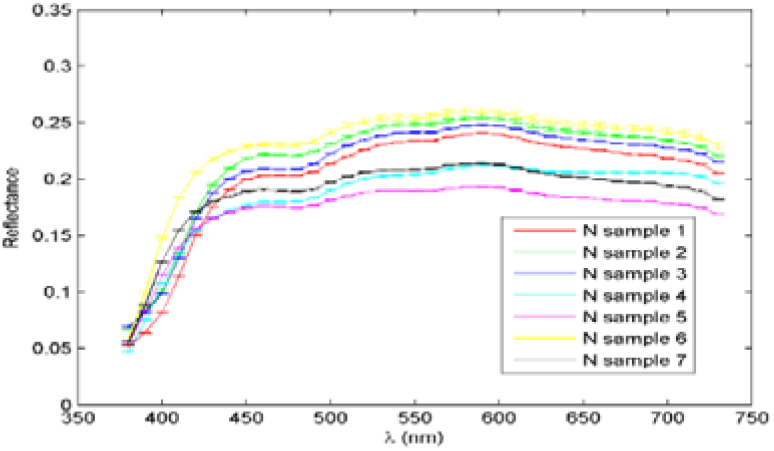
Nano filler Z350 color CIELAB before exposure

**Diagram 2.**
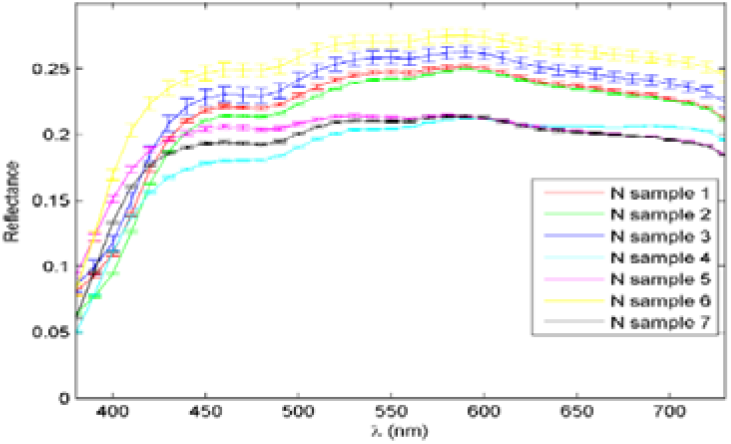
Nano filler Z350 color CIELAB after exposure

**Diagram 3.**
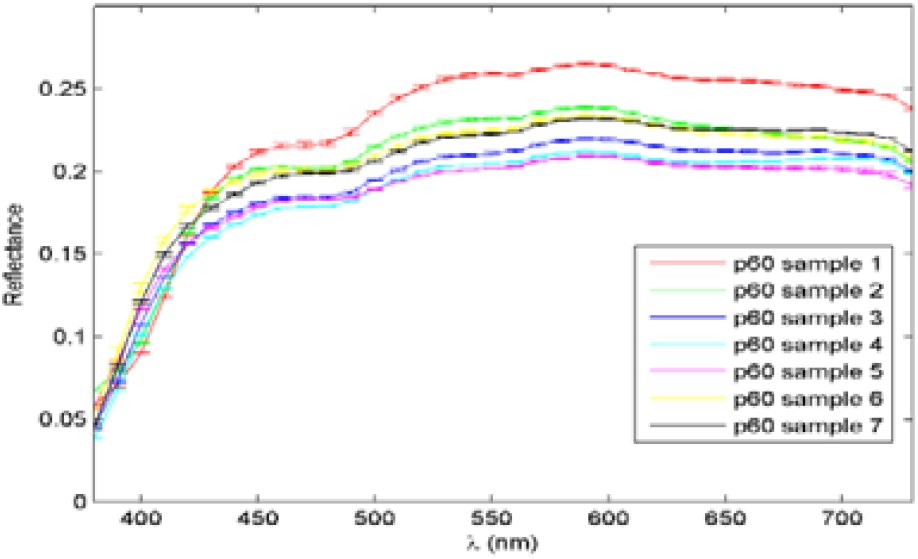
Packable P60 color CIELAB before exposure

**Diagram 4.**
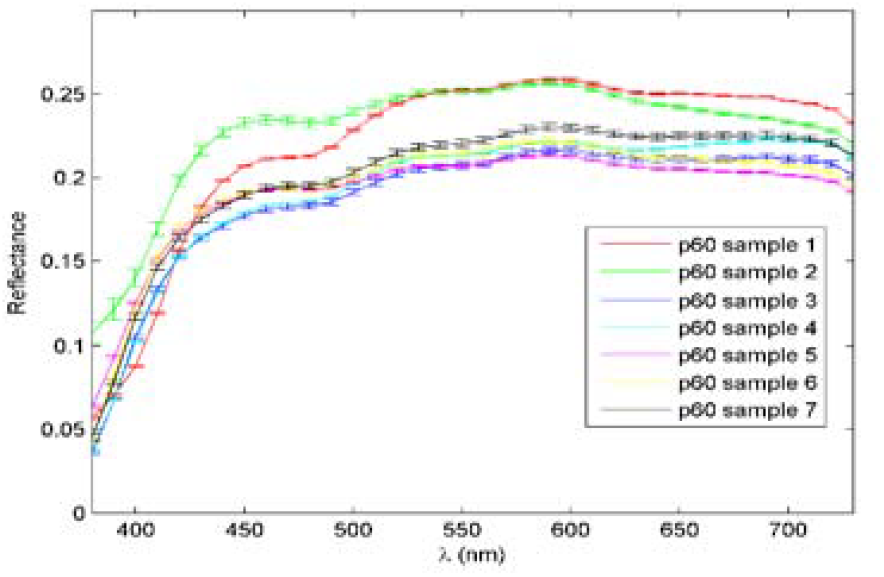
Packable P60 color CIELAB after exposure

**Diagram 5.**
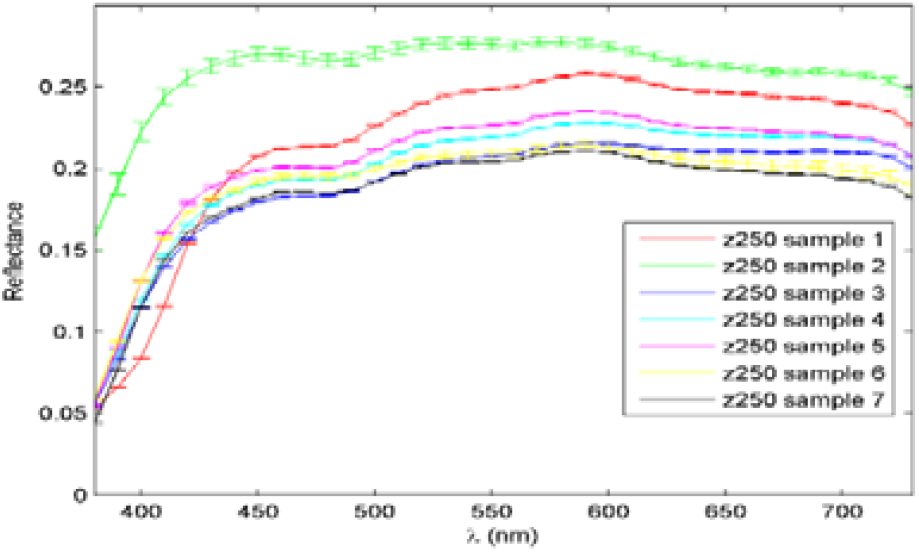
Micro hybrid Z250 color CIELAB before exposure

**Diagram 6.**
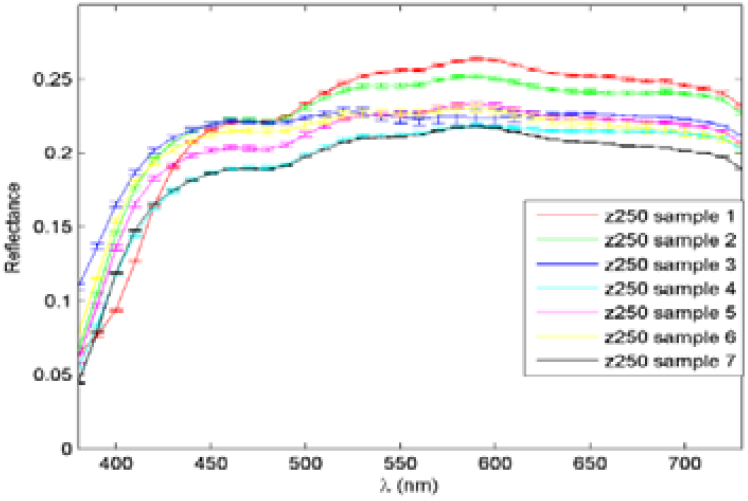
Micro hybrid Z250 color CIELAB after exposure

## Discussion

The present study investigated the effect of chlorhexidine mouthwash on Nanofiller, p60 and z250 composites. The most important side effect of chlorhexidine is tooth discoloration. This discoloration occurs due to the cationic nature of the chlorhexidine molecule and local deposits due to the reaction between chlorhexidine adhering to the tooth surface and chromogens in foods and beverages. Studies have shown that although chlorhexidine is prone to discoloration on natural teeth, it has no significant effect on restorative composites.^13^

We reviewed the results in four different areas. These four areas include the color changes in the green and red spectrum (Δa), color changes in the yellow and blue spectrum (Δb), changes in color transparency and brightness (ΔL)^14^

The combination of all three different areas is similar to what happens in the human eye, and we understand the color of a composite as a combination of three different areas. According to the results obtained from the first area, the average change of packable, Nano filled and micro hybrid composites is 0, +1, and -0.4, respectively, which means that packable composite has no red-green color change and micro hybrid composite underwent the highest color change towards the green spectrum. ^15^

In clinical observations, the z250 micro-hybrid composite shows a to gray color change, which can justify further color change in the z250 spectrum, which can be attributed to the type of fillers.

In yellow-blue color changes, p60 composite underwent the highest blue color changes with one-degree change, followed by micro-hybrid composite with -0.3 change to blue color, and Nano filled composite with the least color change to the blue spectrum (−0.2).

So far, the results are surprisingly similar to the clinical results of chlorhexidine use in composites, and all composites show green-blue changes.^16^

The reason that packable composite has undergone the most changes is improper polishing property due to its large filler particles.

On the other hand, there were positive changes in brightness of all three types of composites, which means that the composites can be attributed to their more transparent property after exposure to chlorhexidine or more light-scattering ability.^17^

Overall, changes above number one are visible to the naked eye, and, therefore, many of these changes are technically expressed and may not be visible to the naked eye. ^18^

In the combination of the previous three criteria, which are determined by ΔE, the highest changes were related to nanocomposites, packable and micro-hybrid composites, respectively.

All numbers are greater than one and the changes are visible to the naked eye. There is no significant difference between the composite types, although the microhybrid composite has undergone the best changes, which can be justified considering the fact that the microhybrid composite is better polished than the packable composite and has less water absorption than the Nano filled composite.

According to the results of the present study and related limitations, it can be concluded that micro hybrid composite is the best type of composite in tooth restoration of people who have to use chlorhexidine for a long time.

This is an in vitro study. It is recommended to perform in vivo studies to obtain more accurate results.

## Conclusion

The combination of all three different areas is similar to what happens in the human eye, and we understand the color of a composite as a combination of three different areas. According to the results obtained from the first area, the average change of packable, nanofilled and micro hybrid composites is 0, +1, and -0.4, respectively, which means that packable composite has no red-green color change and microhybrid composite underwent the highest color change towards the green spectrum. So far, the results are surprisingly similar to the clinical results of chlorhexidine use in composites, and all composites show green-blue changes.^19^ The reason that packable composite has undergone the most changes is improper polishing property due to its large filler particles .^20 21^According to the results of the present study and related limitations, it can be concluded that microhybrid composite is the best type of composite in tooth restoration of people who have to use chlorhexidine for a long time.

## Data Availability

All data produced in the present study are available upon reasonable request to the authors

## Ethical approval

our article have a ethical approval by number of 4013320 in Aja university and don’t have any ethical problem.

